# Leukocyte Telomere Length and Cardiac Structure and Function: A Mendelian Randomization Study

**DOI:** 10.1101/2023.09.13.23295516

**Authors:** Ahmed Salih, Ilaria Boscolo Galazzo, Gloria Menegaz, André Altmann

**Affiliations:** William Harvey Research Institute, NIHR Barts Biomedical Research Centre, Queen Mary University of London, Charterhouse Square, London, EC1M 6BQ, UK; Department of Population Health Sciences, University of Leicester, Leicester, UK; University of Verona, Department of Computer Science, Verona, Italy; Centre for Medical Image Computing (CMIC), Department of Medical Physics and Biomedical Engineering, University College London, London, UK

**Keywords:** Telomere, Cardiac IDPs, Mendelian Randomization

## Abstract

Existing research demonstrates association of shorter telomere length (TL) with increased risk of agerelated health outcomes including cardiovascular diseases. However, the direct causality of these relationships is not definitively established. Cardiovascular aging at an organ-level may be captured using image derived phenotypes (IDPs) of cardiac anatomy and function. In the current study, we use two-sample Mendelian Randomization (MR) to assess the causal link between TL and 54 cardiac magnetic resonance imaging (CMR) measures representing structure and function across the four cardiac chambers. Genetically predicted shorter TL was causally linked to smaller ventricular cavity sizes including left ventricular end-systolic volume (LVESV), left ventricular end-diastolic volume (LVEDV), lower left ventricular mass (LVM) and pulmonary artery. The association with LVM (*β* = 0.217, P_FDR_ = 0.016) remained significant after multiple testing adjustment, whilst other associations were attenuated. Our findings support a causal role for shorter TL and faster cardiac aging, with the most prominent relationship with LVM.

## 1. Introduction

Global population aging has increased the burden of chronic non-communicable diseases of older age, among which, cardiovascular diseases (CVDs) are the most prominent [1]. The risk of age-related CVD is not uniform across the population. Indeed, biological cardiovascular aging is influenced by a wide range of environmental and lifestyle exposures throughout life [2]. Furthermore, the susceptibility to CVDs varies even among individuals with similar exposure profiles, suggesting the crucial role of inherent genetic determinants of biological aging.

Telomeres are distinct regions of repetitive nucleotide sequences and protein complexes at the end of chromosomes [3]. Their function is to protect against nucleotide degradation, unnecessary recombination, repair, and inter-chromosomal fusion [4, 5]. Telomeres become progressively shorter with each cell division.

Shorter telomere lengths (TL) have been linked to increased risk of death and age-related disease, including a range of CVDs [6]. For instance, longer TL are causally associated with decreased risk of myocardial infarction, coronary atherosclerosis and ischemic heart disease [7]. There is wide variation of TL across the population with evidence of high and consistent heritability [8, 9]. As such, TL has been proposed as an indicator of biological aging providing information beyond chronological age and environmental exposures. However, TL can also be modified by diseases, such as diabetes and atherosclerosis, which may confound its relationships with incident CVD and death [6]. That is, the causality of the relationships between TL and CVD is not adequately demonstrated in existing literature [10]. Cardiac magnetic resonance (CMR) became an essential tool for the characterization of CVDs. The cardiovascular phenotype alters in a predictable and established manner with increasing age, which may be accurately captured using CMR [11]. Thus, CMR phenotypes provide reliable continuous measures of cardiovascular aging. The recent availability of genome wide association study (GWAS) data [12, 13, 14, 15] for a rich array of CMR phenotypes provides opportunity to evaluating causal associations of TL with cardiovascular aging.

A statistical genetics technique that can help establish causality from GWAS results is Mendelian Randomization (MR). A recent study used MR to examine the association between TL and seven CMR metrics [16]. The findings indicate that shorter TL was associated with reduced left ventricular mass, reduced left ventricular stroke volume, reduced global ventricular volume and reduced overall ventricular size. However, only the reduction in left ventricular mass would survive multiple testing correction across the seven explored traits. In particular, the study implemented a one-sample MR, where the exposure (TL) and outcome (CMR metrics) are obtained from the same individuals, which enabled the authors to leverage a large dataset comprising N=460,000 individuals to identify instrumental genetic instruments of TL for the MR analysis. However, such analyses are known to be more prone to bias compared to two-sample MR [17]. For instance, ‘weak instruments’ bias the result towards the observed association between exposure and outcome [17], whereas ‘winners curse’ may lead one-sample MR to underestimate the true causal effect [18]. Therefore, a two-sample MR, where there is no overlap between the samples of the exposure and the outcome and which avoids such biases, can be used to substantiate these findings. Finally, Aung et al [16] focused on the most common CMR metrics. However, there are further metrics, such as ejection fraction, which are of broad interest and can be explored using available GWAS summary statistics in a two-sample MR framework.

In this study we used two-sample MR to examine the causal association of TL with 54 CMR phenotypes, leveraging publicly available GWAS summary statistics of TL and cardiac phenotypes. This work provides novel insights into the causal associations of TL with cardiovascular aging and its potential as a marker of biological cardiovascular age, independent of age and environmental exposures.

## 2. Data

### 2.1 Cardiac IDPs GWAS

#### Right atrium

The GWAS summary statistics of the right atrium [12] that is publicly available was considered in the current analysis. Right heart structure and function GWAS was conducted for 40,000 participants from UK Biobank (UKB).

Participants were excluded if they had history with specific heart diseases. GWAS analysis was conducted for 34 CMR measures including right ventricular volume at end diastolic and end systolic, ejection fraction and stroke volume. In addition, Pirruccello et al [12] conducted GWAS for the pulmonary system, including pulmonary root diameter, pulmonary artery (PA) and diameter of PA. Similar measures were also used but indexed for body surface area. Whilst their analysis was mainly focused on right atrium, they also conducted GWAS for left and right ventricular metrics. The full list of all 34 CMR measures reported and the GWAS results can be found in [12].

#### Right ventricular

To represent right ventricular phenotypes, we used publicly available GWAS summary statistics of the following four right ventricular measures: end systolic volume (RVESV), stroke volume (RVSV), ejection fraction (RVEF) and end diastolic volume (RVEDV) [13]. In summary, the results are based on 29,506 participants from UKB free from pre-existing myocardial infarction or heart failure.

#### Left atrium

GWAS was performed using five functional and volumetric left atrium (LA) variables from UKB for 35,658 participants. The variables included LA minimum (LAmin) volumes, indexed LA maximum (LAmax), LA active emptying fraction (LAAEF), LA passive emptying fraction (LAPEF), and LA total emptying fraction (LATEF) [14]. Left ventricular We used the GWAS summary statistics for the following six left ventricular measures: end-diastolic volume (LVEDV), mass (LVM), end-systolic volume (LVESV), ejection fraction (LVEF), mass to end-diastolic volume ratio (LVMVR) and stroke volume (LVSV) [15]. However, as the summary statistics of LVSV were not available, this measure was excluded from further analyses. The study was conducted on 16,923 European participants from UKB.

#### Aortic distensibility

GWAS was conducted to assess the association of the genetic bases and six aortic dimension and distensibility phenotypes. These included ascending aortic distensibility, ascending aortic minimum area, ascending aortic maximum area, descending aortic distensibility, descending aortic minimum area, descending aortic maximum area [19]. The analysis was conducted on 32,590 Caucasian participants from UKB.

#### Arterial stiffness index

Arterial stiffness index (ASI) GWAS was conducted on 127,121 European-ancestry individuals from UKB [20]. Such analysis has been facilitated by its ease of acquisition and its role in a wide range of cardiovascular diseases.

The analysis ended up with a set of variants in four loci that are significantly associated with ASI.

### 2.2 TL GWAS

We used 33 single nucleotide polymorphisms (SNPs) that have been shown to be significantly associated with TL in previous studies [21, 22, 23]. The first 20 SNPs were from a recent study that was conducted on 78,592 European individuals, under the ENGAGE project (European Network for Genetic and Genomic Epidemiology). The remaining 13 SNPs were used before in [24] to conduct MR analysis with a wide range of aging-related outcomes. The focus on these studies, which did not use UKB data, for the selection of the instrumental variables allows us to conduct a two-sample MR study using CMR GWAS summary statistics that were obtained using UKB participants.

The process of choosing proxies for SNPs that are not available for the GWAS on CMR measures and ensure that they are not in linkage disequilibrium (LD) was performed as described previously [25]. Briefly, ten SNPs were excluded as they were in high LD (R^2^ *>* 0.02) with other SNPs. Ensembl [26] was used to calculate LD using GBR (British in England and Scotland) samples from Phase 3 (version 5) of the 1,000 Genomes Project. The final list of the 23 SNPs is shown in Table 1.

**Table 1:**
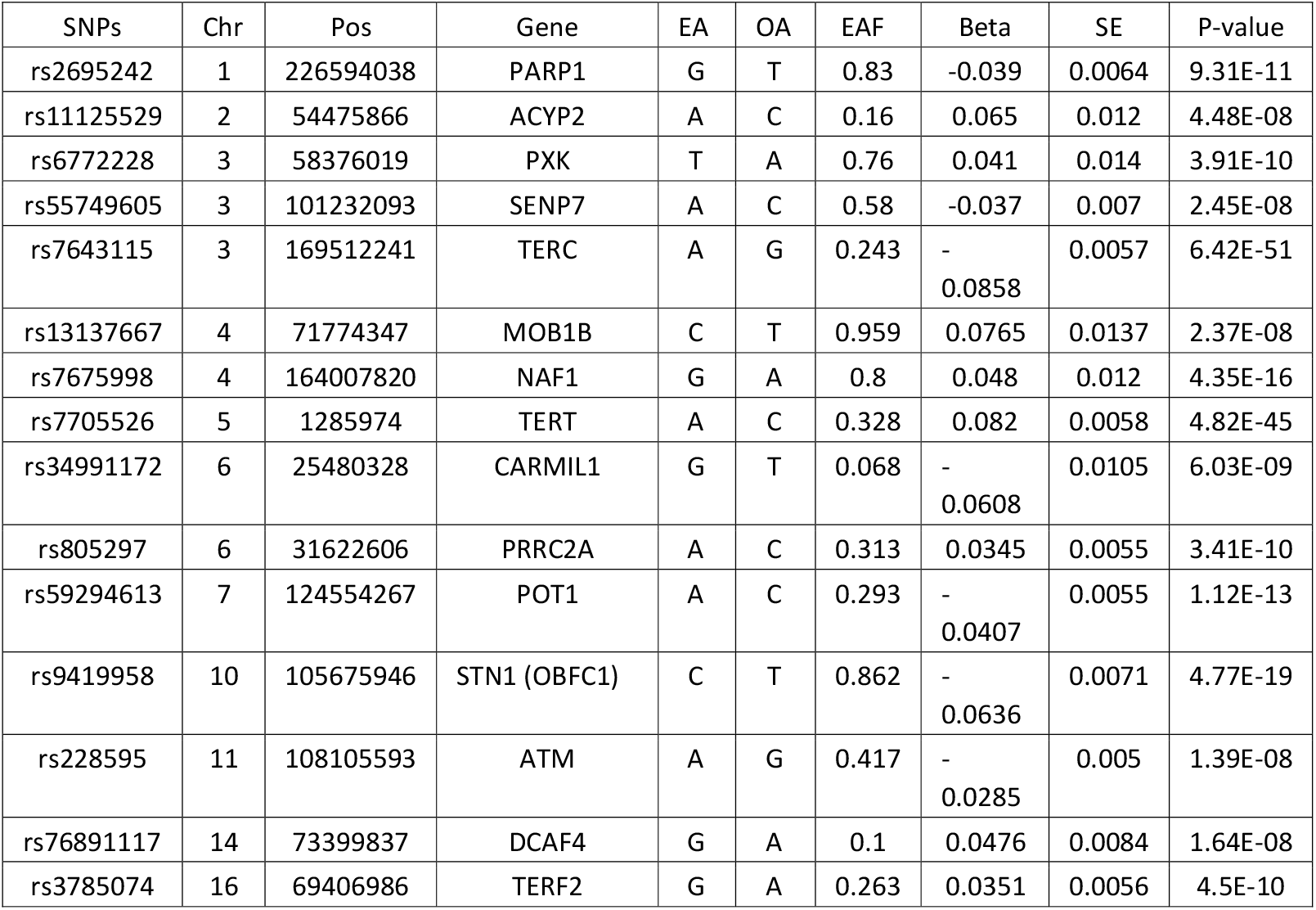
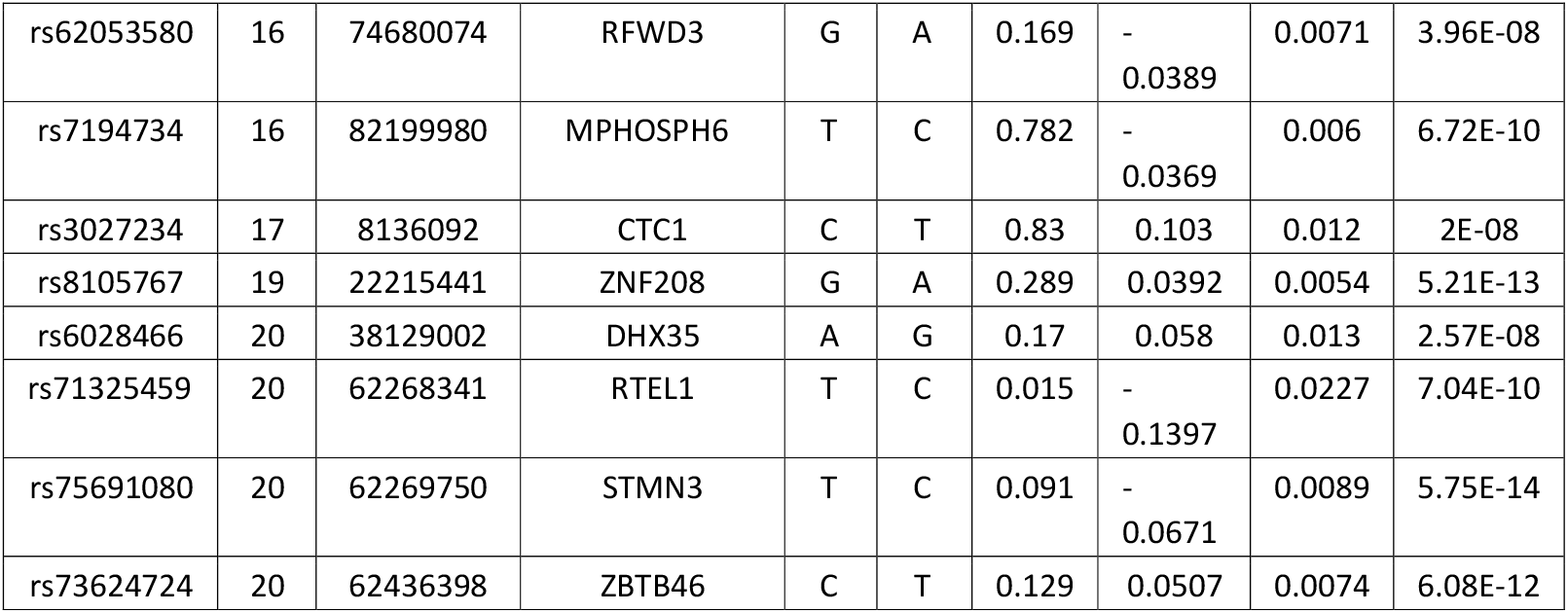
List of the SNPs used in the MR analysis. The SNPs in bold are proxies as the original SNPs were not available for cardiac IDPs GWAS. SNPs, ID of the SNP; Chr, chromosome; Pos, position of the SNP in the genome; EA, effect allele; OA, other allele; EAF, effect allele frequency; Beta, beta value of the SNP in GWAS; SE, standard error.

## 3 Methods

The statistical analysis was implemented using TwoSampleMR package [27] in R adopting inverse variance weighted (IVW) method as the primary analysis. Weighted mode (WMo) and weighted median (WMe) methods were also performed as complementary MR analyses. MR-Egger regression [28] and weighted median function [29] were used to detect heterogeneity and directional pleiotropy of the genetic instruments. To test for horizontal pleiotropy test, leave-one-SNP-out analyses, MR-Egger intercept test, and the modified Cochran Q statistic methods were applied. Moreover, MR–Pleiotropy Residual Sum and Outlier (MR-PRESSO) [30] was used to detect and correct pleiotropy. False discovery rate (FDR) [31] correction was finally applied to the IVW results to adjust for multiple tests across the 54 cardiac GWAS studies independently at *α* = 0.05.

## 4. Results

In total 54 MR tests were conducted to examine the causal association of TL and CMR phenotypes. All the instrumental variables used from TL GWAS were also available for cardiac IDPs, with the exception of two SNPs from the LV GWAS [15]. We could not select suitable proxies for these two SNPs, since all variants in the region showed low linkage disequilibrium (LD < 0.01). Consequently, these two SNPs were excluded when MR was conducted between TL and LV IDPs. Five measures showed nominally significant association with TL in both the main and complementary analyses (Table 2). Specifically, we found significant causal association of shorter TL with smaller LV cavity volumes in end-diastole and end-systole (lower LVEDV and LVESV) and lower LVM. Shorter TL was also significantly associated with smaller short axis PA size (in both indexed and non-indexed versions of this metric). There was no evidence of significant horizontal pleiotropy in these five measures, as per MR-PRESSO. In addition, the MR-Egger intercept p-value was not significant indicating no horizontal pleiotropy. The association with LVESV was not significant in one of the complementary analyses (WMe), whilst significant in other analyses. The associations with short axis PA size (in both indexed and non-indexed versions of this metric), LVEDV and LVESV were attenuated to statistically non-significant after multiple testing adjustment (P_FDR_ > 0.05). The association with LVM was the most robust, remaining statistically significant after multiple testing adjustment (P_FDR_ = 0.016). The remaining CMR measures were not significantly associated with TL (Table S1). The results showed that shortening TL over time causes reduction particularly in LVM and suggestively in four other measures. Figure 1 shows the direction of the association of the five cardiac IDPs and TL. It shows that TL shortening causes reduction of all of these metrics over time.

**Table 2:** Results of the Mendelian randomization for the CMR measures that have significant association with TL. IVW: Inverse variance weighted ; WMe: Weighted median; WMo: Weighted median; se: stander error ;pa: pulmonary artery; i: indexed to body surface area.

| CMR measure | IVW (beta) | IVW (se) | IVW (p-value) | IVW ($P_{FDR}$-value) | MR Egger (p-value) | WMe (p-value) | WMo (p-value) | MR-PRESSO (p-value) | Source |
| --- | --- | --- | --- | --- | --- | --- | --- | --- | --- |
| LVEDV | 0.138 | 0.054 | 0.011 | 0.198 | 0.011 | 0.003 | 0.025 | 0.44 | [15] |
| LVESV | 0.118 | 0.054 | 0.029 | 0.392 | 0.027 | 0.141 | 0.031 | 0.61 | [15] |
| LVM | 0.217 | 0.061 | 0.0003 | 0.016 | 0.048 | 0.005 | 0.029 | 0.27 | [15] |
| short_axis_cm_pa | 0.085 | 0.043 | 0.049 | 0.486 | 0.012 | 0.007 | 0.013 | 0.06 | [12] |
| short_axis_cm_pa_i | 0.103 | 0.038 | 0.007 | 0.189 | 0.006 | 0.005 | 0.014 | 0.22 | [12] |

**Figure 1:**
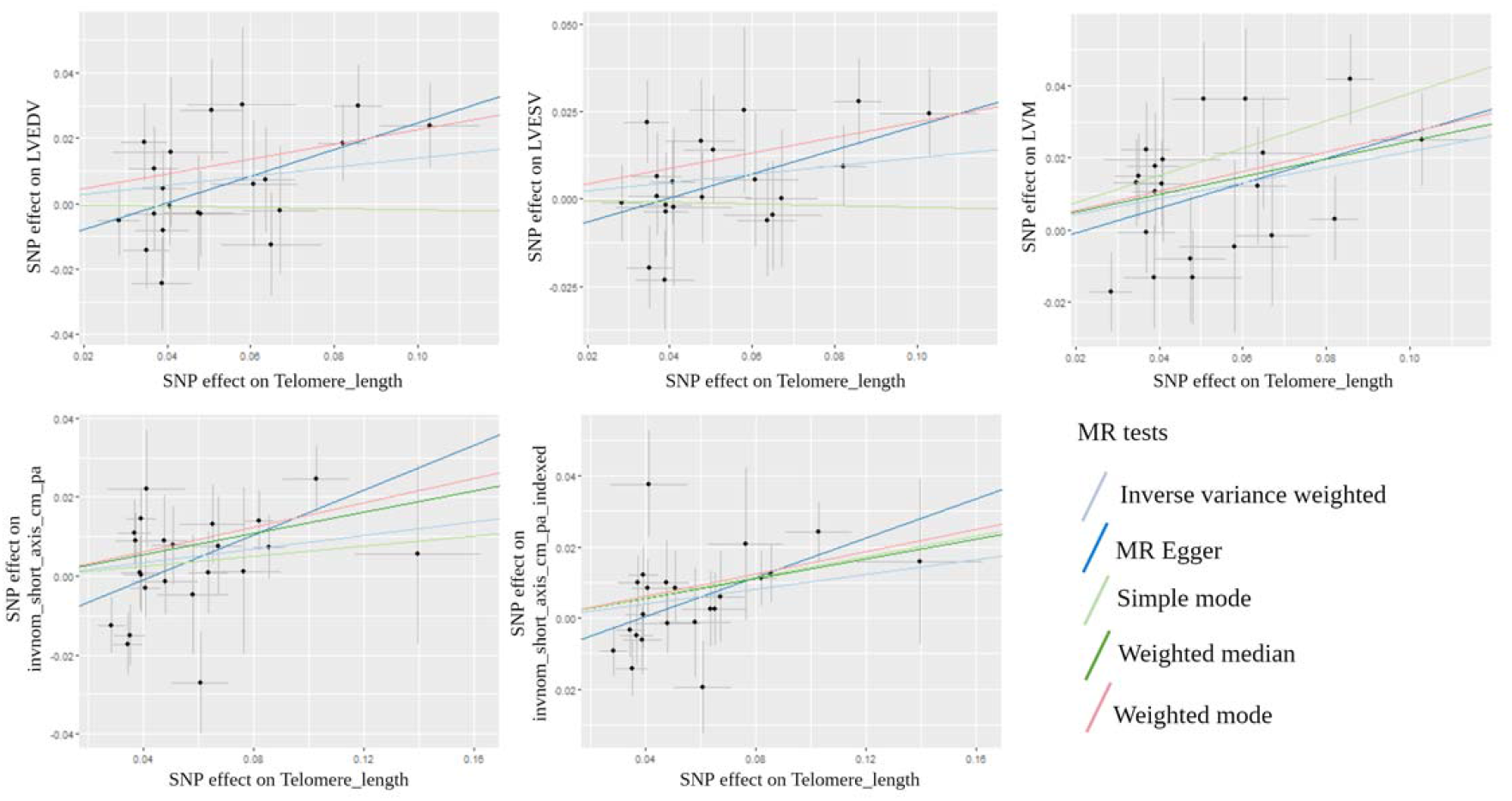
Association of TL and the five cardiac IDPs in the primary (IVW) and in the complementary analyses.

## 5 Discussion

We present an extensive evaluation of the causal relationships between TL and 54 cardiac IDPs using two-sample MR analysis. We demonstrate causal association of shorter TL with smaller LV cavity volumes (smaller LVEDV and LVESV) and lower LVM. The most robust association was between TL and LVM, with the association remaining significant after multiple testing correction of the primary analysis. The pattern of associations reflects associations of TL with a pattern of cardiovascular phenotypic alterations in keeping with greater cardiac aging. These phenotypic alterations have been shown to confer significant cardiovascular risk, particularly when present at younger ages. Thus, our work supports a causal role for TL in driving poorer cardiovascular health and sheds light into potential mechanisms causing differential susceptibility to CVD across the population. The pattern of phenotypic alterations observed in association with shorter TL is consistent with age-related cardiovascular changed reported in population imaging cohorts [32]. There are distinct age-related morphological alterations of the heart, which are reflected in cardiovascular imaging phenotypes and widely reported in the literature [11]. Increasing age is associated with cardio-myocyte attrition [33], which is detected as lower LV mass on CMR.

There is also substantial decline in LV volumes with increasing age, which is disproportionately greater than the LV mass decline, therefore resulting in a concentric pattern of LV remodelling in older ages [32]. With regards to functional metrics, the pattern is more complex, poorer strain metrics are generally noted, although their measurement on CMR can be challenging. LV stroke volume is reduced with increasing age, but accompanied by minimal or no change in LV ejection fraction. Increased vascular stiffness with older age is well described in the literature [34] and linked to the described ventricular remodelling through patterns of ventricular-arterial coupling [35]. The described age-related cardiovascular alterations have been linked to increased risk of cardiovascular events.

Thus, the observed associations with shorter TL in our study are well matched to existing knowledge on cardiac age-related remodelling.

Whilst our analysis indicates causal relationship between shorter TL and age-related LV remodelling, it is not clear whether this represents a direct effect of TL or an indirect effect mediated through greater propensity to risk factors such as diabetes. Indeed, previous work has been linked shorter TL to poorer glycaemic control [36], higher blood pressure [37, 38, 39], and total cholesterol [40]. It is likely that TL acts through multiple direct and indirect pathways to accelerate cardiac aging and increase propensity to disease. Further studies elucidating these precise mechanisms are essential to advance knowledge in this area. The associations with right ventricular and atrial metrics were not significant in our analysis. This may reflect greater technical challenges in deriving these phenotypes and thus greater noise limiting detection of SNPs in GWAS for these metrics. For instance, the RV has an irregular anatomy, and its segmentation is far more challenging than the LV with greater inter-operator variation [41]. It is also possible that the LV reflects more readily age-related alterations than the other chambers. The associations of CMR metrics with aging are not extensively studied due to limited availability of large population cohorts. As the performance automated segmentation tools improve and with greater availability of population datasets our understanding of the genetic architecture of the heart is expected to improve with greater opportunity to understand its alterations with age.

The instrumental variables (SNPs) used in our analysis belong to wide range of genes that regulate TL and are also linked to the development of cardiac diseases and vascular risk factors. rs11125529, rs7675998, rs8105767 and rs7194734 SNPs belong to the *ACYP2, NAF1, ZNF208 and MPHOSPH6* genes, respectively, which were found to be associated with increasing risk of developing coronary heart disease [42, 22, 43]. rs6772228 belongs to the *PXK* gene which is associated with total cholesterol levels [44]. Finally, the *RTEL1* gene plays a protective role against coronary heart disease [45].

Importantly, our study confirms the previously reported causal association between TL and LVM [16] using two-sample MR, which is less susceptible to bias compared to one-sample MR. Similarly to Aung et al. [16], who reported a nominally significant effect between TL and left-ventricular stroke volume, there were also significant associations between TL and Left ventricular end-diastolic and end-systolic volume, but these did not survive multiple testing correction. Interestingly, our results were achieved using only 23 SNPs as instrumental variables, which were previously reported to be significantly associated with TL, as compared to the 130 SNPs by Aung et al [16] based on a UKB GWAS analysis. Of note, the effect sizes in our study were larger than the ones previously reported using one-sample MR. For example, the effect size of LVM in our study is 0.21, while Aung et al [16] report 0.13. Accordingly, the results of the current study provide additional robust evidence for an causal association between TL and CMR metrics.

To the best of our knowledge, this is the first study that attempts to assess the casual link between TL and a wide range of cardiac IDPs. Our results indicate a significant and causal role of TL on the LV structure. A previous study [2] demonstrated the role of wide range of daily-life style and exposures in cardiac aging. Our study identified a new significant player in cardiac aging which might help to better understand the ageing-driven

## Data Availability

This study did not use any raw data, it rather used GWAS summary statistics of several studies that are publicly available

